# Pharmacokinetic bases of the hydroxychloroquine response in COVID-19: implications for therapy and prevention

**DOI:** 10.1101/2020.04.23.20076471

**Authors:** Mohammad Tarek, Andrea Savarino

## Abstract

Chloroquine/hydroxychloroquine has recently been the subject of intense debate in regard to its potential antiviral activity against SARS-Cov-2, the etiological agent of COVID-19. Some report possible curative effects, others do not. In order to shed some light on this rather controversial topic, we used mathematical modelling to simulate possible scenarios of response to hydroxychloroquine in COVID-19 patients. Our computer-aided simulations show that hydroxychloroquine may have an impact on the amplitude of the viral load peak but that viral clearance is not significantly accelerated if the drug is not administered early enough (i.e. when viral loads range from 1 to 1,000 copies/mL). Although some authors had used the trough plasma concentrations or the theoretical drug distribution in the lung to model the effect of chloroquine/hydroxychloroquine on COVID-19, the theoretical drug response based on the trough whole blood concentrations of the drug agreed well with the results of the clinical trials so far reported. Moreover, the effects of chloroquine/hydroxychloroquine could be fully explained when taking into account also the capacity of this drug to raise cell-mediated responses against the productively SARS-Cov-2-infected cells. On the whole, the present study suggests that chloroquine/hydroxychloroquine has a narrow therapeutic window, which overlaps with the highest tolerated doses. These considerations may have implications for development of anti-COVID-19 combination therapies and prevention strategies.

## Introduction

Chloroquine, and its hydroxy-analog hydroxychloroquine, was first proposed in 2003 for treatment of the severe acute respiratory syndrome (SARS) [1] and, due to its promising in-vitro effects [2], was then repurposed as a drug for treatment of COVID-19. As clinical trials in COVID-19 have so far produced mixed results, further study on the interaction of this drug with the virus/host dynamics *in vivo* is necessary in order to provide insight into the optimal timing of administration and association with other interventions. However, after many decades of clinical use of hydroxychloroquine, part of its pharmacokinetics is still unknown, and a number of studies still rely on the drug plasma concentrations [3], which are not an optimal indicator of the biodistribution of this drug, which is a weak base that becomes readily entrapped in intracellular acidic organelles, due to the Henderson-Hasselbach law [1].

A recent advancement likely to shed more light in the interplay of SARS-CoV-2 with antiviral drugs has been the availability of a mathematical model able to simulate the virus/host interplay and matching clinical observations [4]. Similar models developed by some of the authors of that study had proven able to simulate the viral dynamics in HIV/AIDS, providing important therapeutic hints [5, 6]. In the present paper, we use this model to simulate possible scenarios of response to hydroxychloroquine in COVID-19 patients.

## Methods

In the present paper we used the system of differential equations built by Gonçalves et al. [4], as follows.

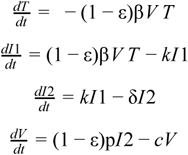

These are ordinary differential equations describing the viral dynamics in a model of target cell population with an eclipse phase accounting for antiviral effectiveness of chloroquine against viral reproduction and target cell populations.

T represents the target cells assumed to be 4×10^8^ [7] based on an assumption of 30 mL nasopharyngeal volume [8] that maintain a cellular concentration of 1.33 × 10^7^ cells/mL [4] which become infected with a constant parameter β which changes according to a constant parameter (1-ε) where ε reflects the antiviral effectiveness of chloroquine simulated at {0%, 33%, 70%,90%} based on an assumption of reduction in the viral basic reproductive number R_0_ by the antiviral [4], I1 represents the infected cells in the eclipse phase, I2 represents the productively infected cells which starts the productivity phase after average time 1/k where k values considered were {1,3,5} d^-1^ to allow prediction of intervention timing effect on viral propagation where releasing virions is controlled by constant parameter p which changes according to a constant parameter (1-ε) and assumed by default to be 10 d^-1^, infected cells are later cleared with a constant parameter rate c which was assumed to be 5 d^-1^ following previous models of viral infection [9], while Infected cells I2 are cleared later from circulation at a constant rate δ assumed by default to be 0.55 d^-1^. V reflects the viral population that was modelled by changing V_0_ at {1^1^, 10^3^, 10^5^} copies/mL (Figure 1 & Supplementary Figures S1, S2). The model neglects the proliferation and death rates of target cells in respect to the concise time scale of infection, instead, the model focuses on the viral effect on target cells.

**Figure 1.**
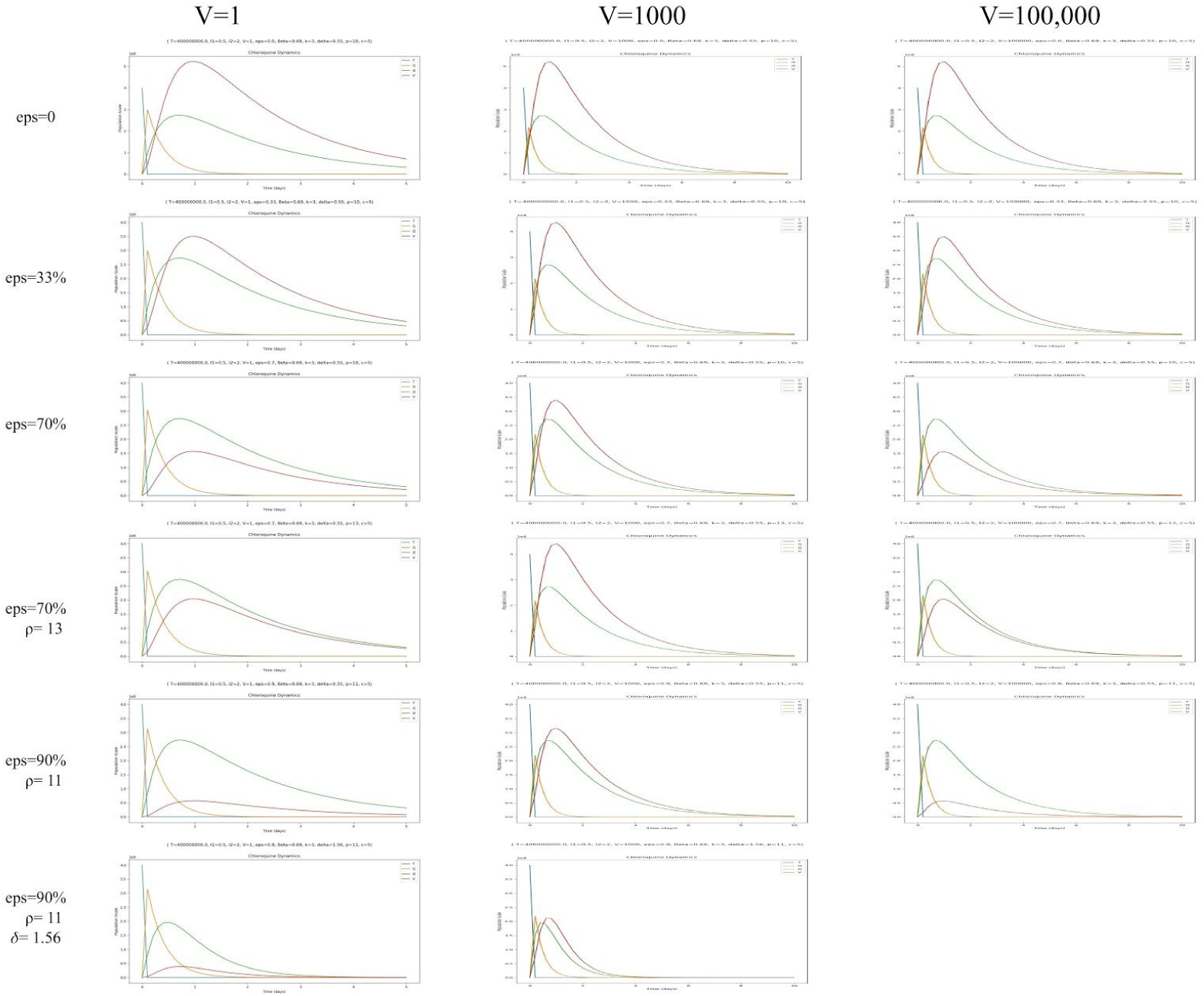
Simulation plots under k = 3 d^-1^ and V_0_ at {1^1^, 10^3^, 10^5^} copies/mL describing the dynamics of reduction in viral load under six conditions; baseline conditions with ε = 0 & p = 10, ε = 33% & p = 10, ε = 70% & p = 10, ε = 70% & p = 13, ε = 90% & p = 11 and ε = 90%, p = 11, δ = 1.56. The lower starting viral loads represent the less advanced stages of COVID-19. The simulation of δ = 1.56 with a starting viral load of 10^5^ copies/mL is not provided as considered to be unrealistic.

Mathematical modelling was implemented using Python programming language v 3.7 (http://www.python.org), ODEINT function of SciPy package was used for solving ordinary differential equations [10], numerical solutions were applied through NumPy package [11] and simulations were plotted within a 10 days’ time frame using Matplotlib package [12].

## Results

Using the system of differential equations developed by Gonçalves et al. [4], we showed that hydroxychloroquine may have an impact on the amplitude of the viral load peak. The effect on viral clearance is only evident in our simulations of mild viral loads, being less pronounced when viral loads are 10^5^ viral RNA copies/mL, i.e. a viral load commonly found in throat swabs during symptomatic disease [13].

Using the parameters adopted by the authors, the model shows that there is an early peak in viral replication followed by a gradual and slow abatement of viral load accompanying consumption of target cells (Figure 1 and Supplemental figures S1 and S2, first row). In their treatment simulations, Gonçalves et al. [4] concluded that hydroxychloroquine had no sufficient effect on the viral load dynamics because the pharmacokinetic parameter that they used to conduct the simulations was the plasma drug concentration, which, as mentioned above, is unlikely to reflect the biodistribution of this drug. In light of the plasma concentrations, the *in-vivo* drug efficacy that they calculated was 33%. The second row of Figure 1 and Supplemental figures S1 and S2 shows our simulations under those conditions. When we used the whole blood concentration (including the drug compartment entrapped in lysosomes) [14], we calculated a drug efficacy of 0.7, based on the concentration-response curves published by Yao et al. [15]. In this case, a more pronounced decrease in the viral peak was observable (Figure 1 and Supplemental figures S1 and S2, third row).

Another aspect that needs attention is the double mechanism of the antiviral effects of hydroxychloroquine. As derived from studies on a huge number of viruses, lysosomal entrapment of hydroxychloroquine is highly likely to affect both viral entry (simulated by Gonçalves et al. [4], and post-entry stages in cellular organelles such as viral budding and viral particle glycosylation [1, 2] (not considered in the aforementioned study). According to the data of Wang et al.[2], chloroquine displayed approximately half of its antiviral activity when it was added after virus adsorption onto cells, confirming that part of its activity occurs at a post-entry level. Therefore, as we also wanted to estimate the impact of hydroxychloroquine-induced impairment of viral production at times beyond the 48 h period considered to estimate hydroxychloroquine efficacy *in vitro* [15], we decreased the viral production rate by 35%, i.e. half of the value calculated on the basis of the drug efficacy on the whole viral replicative cycle. We obtained a further decrease of viral load at peak and steady-state (Figure 1, fourth row).

The data that we used to estimate the response of SARS-CoV-2 to hydroxychloroquine were based on the trough blood concentrations provided by Morita et al. [14], which were based on a drug regimen of 400 mg/day. Gautret et al. [3] recently published the trough serum concentrations of hydroxychloroquine obtained from patients with COVID-19 and treated with a higher dosage (i.e. 600 mg/day) of hydroxychloroquine. Unfortunately, no blood concentrations were reported by Gautret et al., but these could be proportionally derived from the ratios provided by Morita et al. [14], by maintaining a similar plasma/blood concentration ratio. This calculation led to a prediction of drug levels in whole blood of 6.16 μM. By plotting this value to the dose-response curves published by Yao et al. [15], we extrapolated an efficacy of 90%: we adjusted the viral infection and production accordingly and obtained a further decrease in the viral load peak (Figure 1 and Supplemental figures S1 and S2, fifth row). This simulation may also account for hydroxychloroquine administered at 400 mg/day in combination with another antiviral agent with moderate potency.

Among the immune-modulating effects of hydroxychloroquine is its capacity to induce antigen cross-presentation and increase the immune cytotoxic response [16, 17]. Enhanced cytotoxic responses result in increased death of the infected cells. By increasing the death rate of productively infected cells (an effect of cytotoxic immunity) by only 3 fold (data derived from Accapezzato et al. [16]), we could observe a further decrease in the peak amplitude and a shorter time to viral load exhaustion (Figure 1 and Supplemental figures S1 and S2, sixth row).

## Discussion

The simulations that we provide confirm current hypotheses that the virologic response to hydroxychloroquine in COVID-19 patients has a pharmacokinetic basis, and that the drug dosages with an acceptable toxicity profile have a narrow window of overlap with the antiviral effective concentrations. These results are in line with previous analyses of HIV clinical trials, showing that the dosages adopted safely in the clinic are in the lowest range of the therapeutic window, with significant, though yet partial, effects observable only at the highest doses administered [18].

These observations are in line with the clinical data reported so far. A small trial in France suggested that hydroxychloroquine helps treat COVID-19 [3] and a small study from China [19] reported opposite results. Both studies have their own limitations and very limited statistical power, due to the fact that they were prepared during an emergency. The discrepancy of the results obtained may reside, according to the results of the present study, in dosage of hydroxychloroquine adopted, with the French study reporting positive results using 600 mg/day of hydroxychloroquine and the Chinese study being unable to show any efficacy following adoption of a lower dosage (400 mg/day). Furthermore, the reported response to hydroxychloroquine in the French study is incomplete, and the authors had to add another experimental drug repositioned as an antiviral (azithromycin) in order to increase efficacy. These considerations are in line with the results obtained in randomized clinical trials using chloroquine and hydroxychloroquine at an equivalent dosage superior to that reported in the two aforementioned studies [20-22]. As the safety margin of chloroquine/hydroxychloroquine is narrow [23], administration of higher dosages is unfortunately hampered by toxicity, as shown by Borba et al. [22], although recent data show that the toxicity observed by Borba et al. could in fact be ascribed by the concomitant administration of azithromycin [24].

Our results agree in part with the simulations conducted by Arnold and Buckner [25], showing that only the 600 mg/day regimen might have a significant impact on viral replication according to all parameters that they adopted, and those of Garcia-Cremades et al. [26], showing that a dosage higher than or equal to 600 mg/day is necessary to obtain a sustained remission. The approach that we have followed is more conservative than that adopted by Arnold and Buckner [25], in that we used the whole blood concentrations as a measure of prediction of tissue accumulation rather than the predicted accumulation in the lung. According to the model adopted by Arnold and Buckner [25], the drug lung concentrations are hundreds of times higher than those in plasma and are predicted to reach a concentration able to inhibit 100 percent of SARS-CoV-2 replication. We decided not to follow this prediction because 1) tissue accumulation of hydroxychloroquine in lung is mainly driven by tissue macrophages, which are not a main target for the virus, 2) according to the data published by Yao et al. [15], 100 per cent of inhibition of coronavirus replication is not reachable experimentally using hydroxychloroquine. Also Smit et al. reached similar conclusions, based on pharmacokinetic considerations [23].

Based on our simulations, we suggest that chloroquine may not be a “silver bullet” eradicating COVID-19 infection but, on the other side, it might display some beneficial effects *in vivo*. Our simulations go beyond the other models published, indicating that earlier virologic negativization should not be expected when hydroxychloroquine is administered at dosages of 400 mg/day but only at dosages of 600 mg/day, i.e.within the narrow therapeutic window. Instead, we believe that the theoretical capacity of hydroxychloroquine to decrease the viral load peak when viral replication is most active deserves attention and further investigation. The results of our simulations are in line with those of a recent meta analysis of the so far available clinical data, showing that hydroxychloroquine may decrease disease severity but not viral clearance when administered to symptomatic COVID-19 patients [27].

One limitation of the present study resides in the fact that we only modeled the viral dynamics but not the effects of the molecule on immune mediated mechanisms such as the cytokine storm [1]. Be that as it may, an impact on the immune hyperactivation can in this case be inferred from the estimated potential of the drug to decrease the viral load peak, the amplitude of which is associated with deleterious immune hyperactivation in coronavirus infections [1]. This hypothesis is in line with the ability of SARS-CoV-2 to infect CD4+ T-cells [28], the target cells of HIV, which is characterized by immune hyperactivation, as well [18].

Finally, we show that the effect of chloroquine/hydroxychloroquine on antigen cross-presentation should be considered. Cross-presentation is a phenomenon in which a dendritic cell presents the antigen to CD8+ T-cells, improving their priming. Activation of CD8+ T-cells after priming through antigen recognition can induce the selective killing of the infected cells [16]. Taking into account this parameter it is possible to predict that hydroxychloroquine may accelerate viral clearance but only when administered early. Accordingly, we did not model a scenario of increased cell-mediated responses when viral loads are higher, because immune dysregulation is in this case likely to have taken place. Enhanced cell-mediated responses may not only induce selective killing of the infected cells but also enhance immunity for better long-term control of the infection.

Hydroxychloroquine is currently being investigated also as a prophylactic agent [29]. Given the great interest that this strategy has so far evoked in the context of COVID-19 [30, 31], future vaccines against this disease should be considered in co-administration with chloroquine/hydroxychloroquine. In case chloroquine/hydroxychloroquine should prove ineffective in protecting from SARS-CoV-2 acquisition, it is plausible that it may mitigate disease severity by attenuating viral replication.

## Data Availability

The mathematical simulations in this study has not incorporated any clinical from patients and rather focused on pharmacokinetic parameters from previous studies and current clinical trials on chloroquine/hydroxychloroquine for COVID-19.

## Supplementary Material

**Figure S1**. Shows the simulation plots under k = 1 d^-1^ and V_0_ at {1^1^, 10^3^, 10^5^} copies/mL describing the dynamics of reduction in viral load under six conditions; baseline conditions with ε = 0 & p = 10, ε = 33% & p = 10, ε = 70% & p = 10, ε = 70% & p = 13, ε = 90% & p = 11 and ε = 90%, p = 11, δ = 1.56.

**Figure S2**. Shows the simulation plots under k = 5 d^-1^ and V_0_ at {1^1^, 10^3^, 10^5^} copies/mL describing the dynamics of reduction in viral load under six conditions; baseline conditions with ε = 0 & p = 10, ε = 33% & p = 10, ε = 70% & p = 10, ε = 70% & p = 13, ε = 90% & p = 11 and ε = 90%, p = 11, δ = 1.56.

## Notes

### Competing Interest Statement

The authors have declared no competing interest.

### Funding Statement

No external funding was received for this study.

